# A novel truncating variant c.1222DupC in *RBM20* causes cardiomyopathy through haploinsufficiency

**DOI:** 10.1101/2025.09.16.25335490

**Authors:** Priyanka Pant, Yong Huang, Zakiya Ghouse, Fang Bai, Elena Kemmling, Laura Konrad, Rebecca Kistler, Timon Seeger, Michael Gotthardt, Victoria N. Parikh, Maarten M.G. van den Hoogenhof

**Author notes:** Corresponding author: Dr. Maarten MG van den Hoogenhof, PhD, Institute of Experimental Cardiology, Medical Faculty Heidelberg, Heidelberg University, Eppelheimer Strasse 8, 69115, Heidelberg, Germany.

## Abstract

RBM20 is a cardiac splicing factor responsible for splicing of several cardiac genes such as TTN, TRDN, RyR2, PDLIM1, and CAMK2D. Mutations in RBM20 are a major cause of familial dilated cardiomyopathy (DCM), and lead to missplicing of RBM20 target genes. Here, we describe a novel heterozygous truncating mutation, *RBM20* c.1222DupC, identified in a patient with mitral valve prolapse and late onset familial DCM. This mutation introduces a premature termination codon and generates a truncated protein of ∼55 kDa *in vitro*. Splicing assays demonstrated complete loss of activity and no dominant-negative effect on wild-type RBM20. Overexpression in NRCMs revealed that the truncated protein localized to both cytoplasm and nucleus, partially co-localizing with wild-type RBM20, despite lacking the RS and RRM domains. To model the patient’s condition, we generated a heterozygous c.1222DupC mutant induced pluripotent stem cell line and differentiated these in cardiomyocytes. Western blot analysis of endogenous RBM20 revealed a strong reduction in RBM20 protein level. RT-PCR revealed splicing defects in canonical RBM20 targets, and RNA-sequencing identified widespread splicing abnormalities, including in established RBM20 targets (TTN, RyR2, CAMK2D, and CACNA1G). Together, these findings establish RBM20 c.1222DupC as a truncating variant that causes DCM primarily through haploinsufficiency.

## INTRODUCTION

Missense mutations in *RBM20* are an established cause of dilated cardiomyopathy (DCM) and account for approximately 3 to 6% of familial DCM cases^1^. RBM20 regulates the splicing of multiple cardiac genes, including titin (TTN), ryanodine receptor 2 (RYR2), LIM domain binding 3 (LDB3), and calcium/calmodulin-dependent protein kinase II delta (CAMK2D)^1^. RBM20 contains several functional regions: a leucine/proline-rich region, two zinc finger domains, an RNA recognition motif (RRM), an arginine/serine-rich (RS) domain, and a glutamic acid-rich region. Although variants occur throughout the gene, a hotspot in the RS domain in exon 9 is associated with early onset and severe disease, frequently with arrhythmia^1,2^. While wildtype (WT) RBM20 typically exhibits a characteristic bipunctate nuclear pattern, RBM20 with RS-domain missense variants mislocalizes to the cytoplasm, where it forms ribonucleoprotein condensates or aggregates. RS-domain variants include p.P633L (c.1898C>T) and p.R634Q (c.1901G>A) or p.R634W (c.1900C>T), which are associated with severe DCM, and p.R634L (c.1901G>T), which has been reported with left ventricular non-compaction (LVNC)^3–6^. Both P633L and R634Q mislocalize RBM20, while P633L only produces partial mislocalization^4,7^. Additional RS-domain variants, including p.S635A (c.1903T>G), p.R636C (c.1906C>T), and p.P638L (c.1913C>T), also show cytoplasmic mislocalization along with the reported early onset of the DCM phenotype^8,9^. These mutations disrupt the interaction of RBM20 with transportin 3 (TNPO3), its nuclear importer, leading to its mislocalization^7^. Outside the RS domain, variants in the glutamic acid-rich region such as p.E913K (c.2737G>A), p.V914A (c.2741T>C), and p.L908P (c.2723T>C) are associated with DCM without cytoplasmic mislocalization^8,10,11^. Mechanistically, the function of the E-rich domain is not quite clear, although decreased stability of RBM20-E913K suggest that this region could be essential for maintaining the protein stability^11^. The p.I536T (c.1607T>C) variant in the RRM domain, which was identified in a patient with sudden cardiac death, impairs splicing *in vitro*. However, *Rbm20*^I538T^ knock-in mice do not exhibit early sudden death, which suggests that loss of splicing alone may be insufficient to reproduce the early and severe human phenotype^12,13^. Recent work indicates that truncating *RBM20* variants are also associated with arrhythmogenic DCM, although on average they show lower penetrance and a milder clinical course than canonical RS-domain missense variants^14^. Overall, these data suggest that while pathogenic RBM20 mutations all lead to cardiac disease, RS-domain mutations lead to a more severe phenotype. The molecular mechanism(s) underlying RBM20 cardiomyopathy are not entirely clear, but at least two partially distinct mechanisms of RBM20-mediated disease have been proposed. The first is the loss-of-function that leads to missplicing of key cardiac genes, which happens with all disease-causing variants. The second is the mislocalization of RBM20 with condensate formation, which seems to be an additional toxic gain-of-function, that occurs with RS-domain variants. Since patients with these variants would have both missplicing of RBM20 targets and mislocalization of RBM20, this aligns with the heightened arrhythmia risk and DCM severity observed in these patients^15^. Here, we report a novel truncating variant in *RBM20*, a duplication (c.1222DupC in exon 2) which leads to a frameshift and premature stop codon, identified in a patient with mitral valve prolapse and regurgitation who developed persistent DCM after mitral valve repair. Additionally, family history included multiple early deaths, suspicious for sudden arrhythmic cause. In functional studies, we show that the truncated protein is less stable, lacks splicing activity, has no dominant negative effect, and shows mixed nuclear and cytoplasmic distribution. These findings support haploinsufficiency as the disease mechanism for this RBM20 variant.

## MATERIALS AND METHODS

### Patient data

All clinical data were collected under IRB-approved waiver of consent.

### Cloning

Mouse and human RBM20 were cloned from mouse and human cDNA respectively. Flag-tagged RBM20 constructs were created by PCR using Q5 polymerase (NEB) with primers listed in Supplementary Table 1. Ligation was performed using T4 DNA ligase (NEB). Constructs were Sanger sequenced and used as template for introducing the DupC mutation. Mutagenesis was performed using QuikChange XL Site-Directed Mutagenesis Kit (Agilent) according to manufacturer’s instructions using the primers given in Supplementary Table 1.

### Prime editing of hiPSC line

To introduce the pathogenic P408fs^*^8 (c.1222DupC) variant into RBM20 gene, we used the PE2 prime editor and PE3 nickase system as previously described^16^. pegRNA and PE3 nicking sgRNA were designed using pegFinder (http://pegfinder.sidichenlab.org), and oligo sequences are listed in Supplemental Table 1. hiPSCs from a healthy individual were electroporated with a 1:1:1 DNA mixture of PE2-GFP, PE2-pegRNA, and PE3-nicking-sgRNA using the Neon Transfection System (1100⍰V, 30⍰ms, 1 pulse). The following day, GFP-positive cells were sorted via FACS (BD FACSymphony S6) and single-cell cloned into 96-well plates containing CloneR (Stemcell Technologies). Eleven days post-sorting, colonies were picked, genomic DNA was extracted using QuickExtract (Biosearch Technologies), and genotyping was performed using the primers listed in Supplemental Table 1.

### hiPSC differentiation

Differentiation towards cardiomyocytes was carried out following a small molecule Wnt-activation/inhibition protocol. Media was changed to RPMI-B27 without insulin (Life Technologies) supplemented with 4-6 μM CHIR99021 (TargetMol). 72h later, medium was switched to RPMI-B27 without insulin supplemented with 3 μM IWP (Selleck Chemicals). After 48h the medium was switched to RPMI supplemented with B27 with chemically defined media minus insulin for two days, then replaced by RPMI-B27 with insulin (Life Technologies) and refreshed every two days. Beating human iPSC-CMs were observed from day 8-10 post-differentiation. On day 13 post-differentiation, cells were transiently cultured in RPMI without D-glucose (Life Technologies) and with B27 minus insulin and supplemented with 0.2% lactate (Sigma) for 96h for metabolic selection of iPSC-CM populations. Fully differentiated iPSC-CMs were maintained with RPMI medium supplemented with B27. All experiments were conducted with iPSC-CMs between 35 and 45 days after differentiation.

### Isolation of NRCMs

1-2 days old Wistar rat pups were used for the isolation of ventricular cardiomyocytes using the Neonatal heart dissociation kit (130-098-373, Miltenyi Biotech), and Neonatal cardiomyocyte isolation kit (130-105-420, Miltenyi Biotech) as per manufacturer’s information. The cells were counted and plated on the Laminin coated coverslips.

### Transfection of HEK293T cells

HEK293T cells were grown in DMEM media with 10% FBS in a 5% CO2 incubator. GeneJammer transfection reagent (Agilent) was used for transfection, and 1ug of pcDNA-FLAG-RBM20 or pcDNA-FLAG-DupC was transfected per well in a 6-well plate. Empty pcDNA vector was used as negative control.

### Splicing reporter assays

HEK293 cells were plated in 96-well plates and transfected at 50% confluency using PEI40 at a DNA-to-PEI40 ratio of 1:3, with a total of 200 ng plasmid DNA. The DNA mixture consisted of 1 ng of the TTN-IG Ex241-243 splice reporter and a 20-fold molar excess of RBM20 expression plasmids or the control plasmid pcDNA3.1 (Invitrogen, Cat# V79520). Plasmids and PEI40 were pre-incubated for 15 minutes in serum-free medium before being added to the cells. Each transfection was repeated ten times. After 60 hours, cell viability was assessed using PrestoBlue (Thermo Fisher Scientific, Cat# A13261). At the same time point, luciferase activity was measured with the Dual-Luciferase^®^ Reporter Assay System (Promega) on an Infinite^®^ M200 Pro plate reader (TECAN). Firefly luciferase activity was normalized to Renilla luciferase and expressed relative to WT RBM20-transfected cells. Data are presented as mean ± SEM from biological replicates (n = 8). Statistical significance was determined by one-way ANOVA with Bonferroni post-test.

### In vitro transcription

IVT was performed using T3 Messenger Max kit (Thermo Fisher). pT3Ts containing clones were digested with XbaI to linearize the plasmid. Linear plasmids were eluted from gel and 300ng of DNA was taken as template to perform in vitro transcription as per the manufacturer’s protocol. The transcribed RNAs were polyadenylated with E Coli. Poly A polymerase (NEB, M0276L). The polyadenylated RNA was precipitated using LiCl assisted precipitation as per the instructions provided by the manufacturer.

### Transfection of NRCMs

Lipofectamine MessengerMAX mRNA Transfection Reagent (Thermo Fisher) was used to transfect 500 ng of RNA per well in a 24 well plate. Lipofectamine and RNA mix was prepared in Optimem medium per the manufacturer’s instruction. The cells were incubated with RNA-lipofectamine mix for 24h. mRNA of GFP was used as positive control for transfection. Cells were collected for downstream analysis after 24h.

### Immunocytochemistry

Cells were fixed in 4% paraformaldehyde for 15 minutes, washed 3 times in PBS and permeabilized in 0.1%Triton-X/PBS for 10 minutes. Cells were blocked in 4% normal goat serum (NGS) for 1 hour at room temperature (RT) and then incubated with primary antibody in 4% NGS overnight at 4ºC. Secondary antibody incubation occurred in 4% NGS for 1 hour at RT. Nuclear staining was performed as a last step using DAPI. Coverslips were then mounted on glass slides with Vectashield hardset mounting medium (H-1400-10) and images were captured using confocal microscopy (Leica Mica). Primary antibodies used were: rabbit anti-FLAG (1:250, Sigma F7425), mouse anti-FLAG (Sigma F1804-1MG), rabbit anti-RBM20 (1:250, Sigma HPA0377703), rabbit anti-Myc (1:200, Cell Signalling, 2278S), mouse anti-Actinin (1:400, Sigma A7811). Alexa Fluor^®^ 488, Alexa Fluor^®^ 594, Alexa Fluor^®^ 647 conjugated antibodies (1:250, Invitrogen) were used as secondary antibodies.

### Western blotting

Cells were lysed in ice-cold RIPA-buffer (50mM Tris-HCl, 150mM NaCl, 1% NP-40, 0.2% sodium deoxycholate, 0.1% SDS) supplemented with DTT, PMSF, and protease inhibitor cocktail (Thermo). Cell lysates were cleared by centrifuging at 14.000xg for 10 min at 4 ºC. Western blotting was performed according to standard protocols. Briefly, protein concentrations were determined using the BCA protein assay (Pierce) and proteins were resolved by SDS-PAGE and transferred to Polyvindyline difluoride (PVDF) membranes (Merck). PVDF membranes were incubated overnight at 4ºC with the following primary antibodies: rabbit anti-FLAG (1:1000, Thermo scientific), rabbit anti-RBM20 (1:1000, HPA0377703, Sigma) mouse anti-GAPDH (1:5000, MAB374, Sigma). Horseradish peroxidase-conjugated secondary antibodies (Biozol) were used for detection and were incubated for 1 hour at room temperature. Western blots were developed with ECL prime western blotting detection reagent (SantaCruz) and images were acquired using the Vilber Fusion FX (Vilber). Densitometric analysis of Western blots was performed using Image J software.

### RNA isolation and q(RT)-PCR

RNA was isolated using TRIzol (Invitrogen) according to the manufacturer’s protocol. Subsequently, 1 μg RNA was used for cDNA synthesis using reverse transcriptase (Steinbrenner Laborsysteme GmbH). RT-PCR was done using Taq-polymerase using primers in Supplementary table 2. qPCR was performed on a Lightcycler 480 (Roche) using SYBR green premix (Applied biosystems). Analysis of qPCR data was performed using LinRegPCR analysis software^17^. Primers used for qPCR are provided in Supplemental Table 2. Gene expression was normalized to the geometric mean of Gapdh and Hprt expression.

### RNA sequencing

Total RNA was isolated with TRIzol (Life Technologies) according to manufacturer’s instructions. RNA integrity was verified with the Agilent Bioanalyzer (Agilent Biotechnologies) before RNA-seq libraries were prepared using the SMART-Seq Total RNA Pico Input kit (634354, Takara Bio) with the Unique Dual Index Kit (634752, Takara Bio). Libraries were then sequenced on an Aviti sequencer (Element Biosciences). Sequencing reads were mapped to the GENCODE GRCh38 reference genome (release 45, Ensembl 111) using RNA STAR (v2.7.11b) on the Galaxy platform (https://galaxyproject.org)^18^. Gene-level counts were generated with featureCounts (v2.1.1). Raw sequence files have been deposited at GEO (…………). Differential gene expression between DupC and WT groups was analyzed in R using DESeq2 (v1.42.1)^19^. Multiple testing correction was applied using the Benjamini– Hochberg procedure. Genes were considered significantly differentially expressed if they exhibited a (|log2FC| > 0.5) with an adjusted p-value <0.05. Gene ontology analysis was subsequently performed using Cluster profiler (v4.14.6)^20^. To investigate alternative splicing differences, we applied rMATS-turbo (turbo_v4_1_2) on a Linux environment^21^. Five classes of splicing events were examined: exon skipping (SE), mutually exclusive exons (MXE), alternative 3⍰ splice sites (A3SS), alternative 5⍰ splice sites (A5SS), and retained introns (RI). Among these, SE events identified using both junction and exon counts (JCEC) were selected for downstream analysis. Exon inclusion differences between conditions were quantified using ΔPSI (percent spliced-in), calculated as ΔPSI = PSI(DupC) – PSI(WT). SE events were considered significant when the pvalue was less than 0.01 and ΔPSI was greater than 0.1. Data visualization and statistical plots were generated in R using the ggplot2, pheatmap, dplyr, and tidyr packages.

### Statistics

Graphpad Prism was used for data analysis and statistics. Data are presented as mean ± sem, and were analyzed with appropriate statistical tests, as indicated in the respective figure legends. A value of p < 0.05 was considered statistically significant.

## RESULTS

### Identification of a novel RBM20 variant

A male patient in early 50s presented with mitral valve (MV) prolapse and severe mitral regurgitation, and developed DCM immediately after MV repair. Family history showed multiple family members with DCM and sudden cardiac death (Figure 1A). Targeted clinical cardiomyopathy and arrhythmia gene-panel sequencing of the proband identified a novel truncating mutation in RBM20, which constituted the insertion of a cytosine at position c1222. This insertion leads to a frameshift and a premature stop codon 8 amino acids downstream of the mutation site (p.Leu408ProfsX8) (Figure 1B). The patient experienced multiple runs of nonsustained ventricular tachycardia (VT) lasting up to 24 beats. Cardiac MRI showed a left ventricular ejection fraction of 39%, moderate left ventricular enlargement, and delayed gadolinium enhancement of both papillary muscle and the mesocardium of the lateral wall. A primary prevention ICD was implanted at that time and the patient was started on guideline directed medical therapy for heart failure. No additional major arrhythmias or heart failure hospitalizations were recorded since diagnosis.

**Figure 1.**
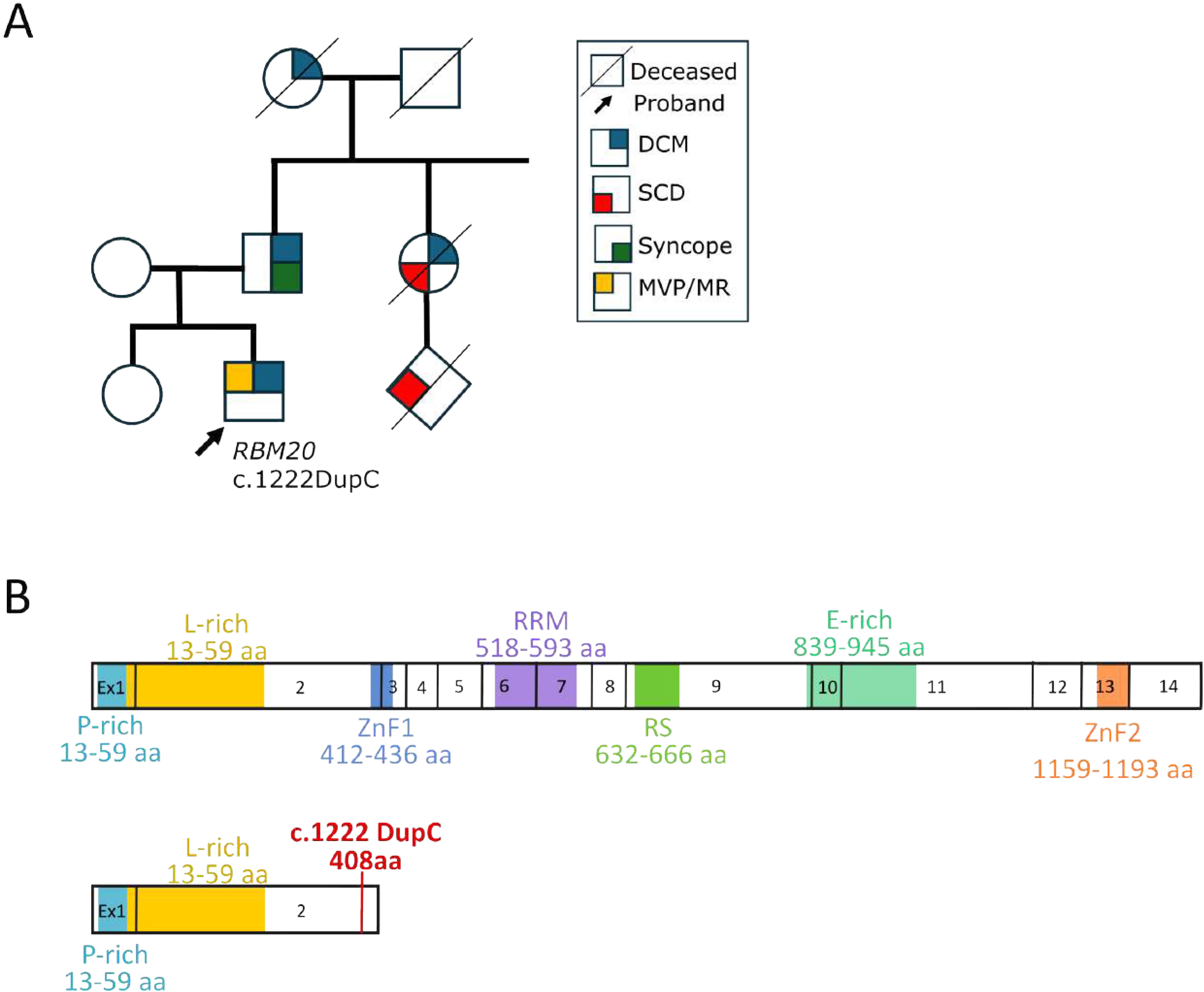
Identification of the *RBM20* c.1222DupC mutation. **(A)** Partial pedigree of the proband with the *RBM20* c.1222DupC mutation. DCM, dilated cardiomyopathy, SCD, sudden death, MVP, mitral valve prolapse, MR, mitral regurgitation. Proband is marked by arrow. **(B)** Schematic visualization of RBM20 protein structure, numbers represent exons and known functional domains are denoted in different colors. Top diagram represents the full length RBM20 and bottom diagram represents truncated RBM20 c.1222DupC protein with the position of the mutation marked with a red vertical line.

### Protein stability of the truncated RBM20-DupC protein is decreased as compared to the WT RBM20 protein

To determine if the *RBM20*-c.1222DupC mutation has an effect on translation, we generated FLAG-tagged constructs of mouse and human WT and DupC mutant *RBM20*, and validated the mutation using Sanger Sequencing (Figure 2A). We then transfected FLAG-tagged human and mouse WT *RBM20* and the DupC mutant in HEK293 cells. We measured the transcript levels of both the WT and DupC *RBM20* and found no significant difference (Figure 2B). Immunoblotting revealed that the DupC mutant produced a protein of approximately 55 kDa, whereas the full-length WT-RBM20 showed its typical size of 180 kDa (Figure 2C). Densitometric analysis revealed ∼50% reduction for mouse and ∼70% reduction for human truncated RBM20 compared to the respective full-length protein, suggesting a decrease in protein stability (Figure 2D).

**Figure 2.**
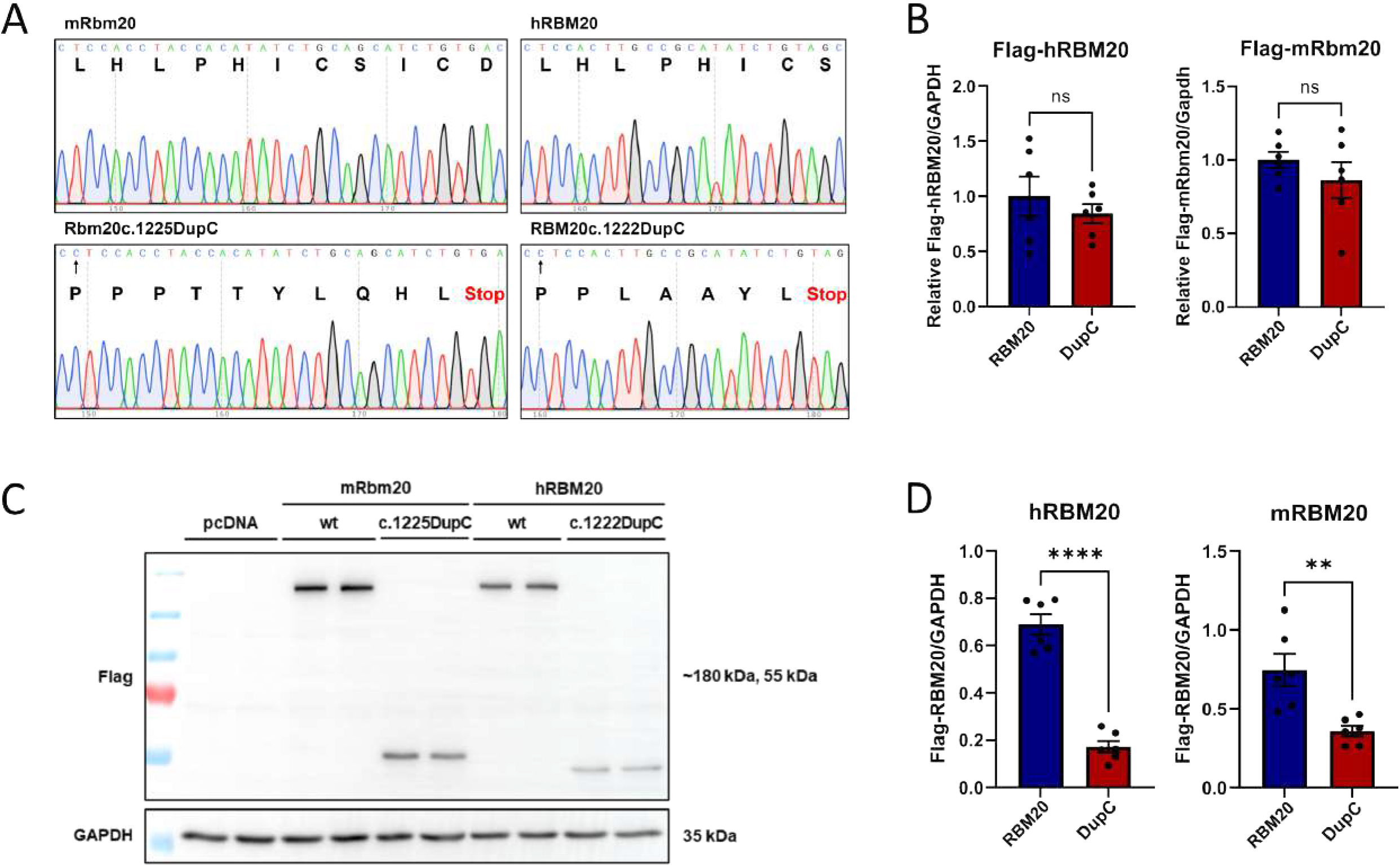
The truncated RBM20-DupC protein is less stable. **(A)** Sanger sequencing of the mouse (left) and human (right) WT (top) and DupC mutant (bottom) RBM20 constructs. Amino acid sequence is added underneath the nucleotide sequence. The arrow in the bottom panels indicates the frame shift after the c.1222DupC mutation. Premature stop codon indicated in red. **(B)** Expression of mouse and human RBM20 mRNA transcript in HEK293 cells transfected with equal amounts of plasmid with mouse or human WT-RBM20 or RBM20-DupC. **(C)** Immunoblotting of FLAG-tagged full length RBM20 and RBM20-DupC in HEK293 cells. **(D)** Densitometric analysis of the FLAG-tagged RBM20 and FLAG-tagged RBM20-DupC normalized to GAPDH intensity. ^**^ *p* value < 0.01, ^****^ *p* value <0.0001. Significance was tested using a two-tailed t-test.

### RBM20-DupC shows both nuclear and cytoplasmic localization

To assess the localization of the RBM20-DupC, we overexpressed FLAG-tagged human and mouse WT RBM20 and the DupC mutant in neonatal rat cardiomyocytes (NRCMs). Both human and mouse WT RBM20 demonstrated nuclear localization with the characteristic bi-punctate pattern (Figure 3A). In contrast, DupC mutants showed both nuclear and cytoplasmic localization. Interestingly, a portion of the nuclear-localized DupC protein appeared to co-localize with the WT RBM20. To further confirm this co-localization, we co-transfected NRCMs with FLAG-tagged human DupC and Myc-tagged human WT-RBM20. Confocal imaging revealed that RBM20-DupC was indeed present in both the cytoplasm and nucleus, and within the nucleus, it co-localized with WT RBM20 (Supplementary Figure 1).

**Figure 3.**
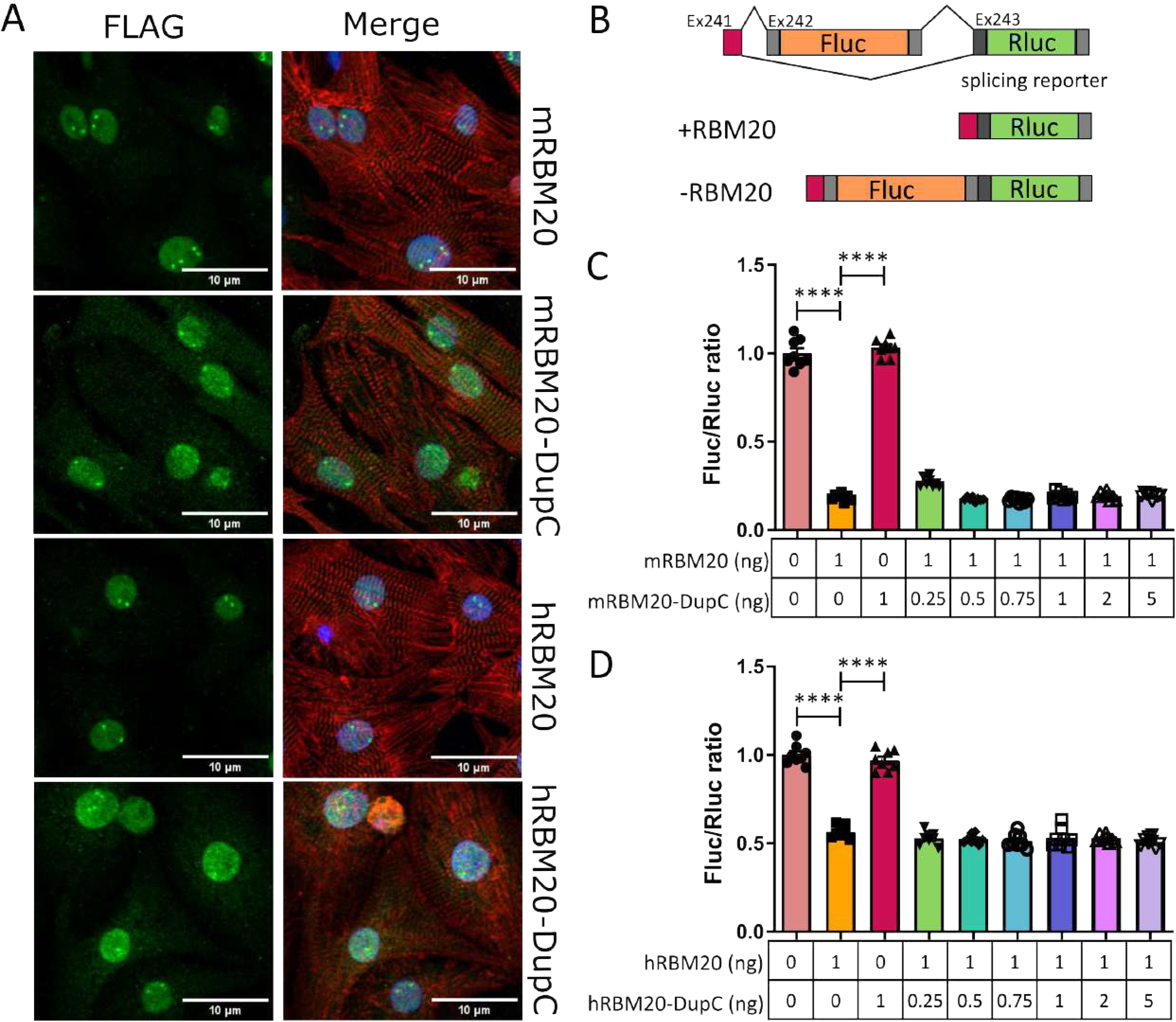
RBM20-DupC does not have residual splicing activity and does not act as a dominant negative. **(A)** Immunofluorescence of NRCMs transfected with mouse or human FLAG-tagged RBM20 or FLAG-tagged RBM20-DupC. FLAG is stained in green, α-actinin in red, and DAPI in blue. Scale bar is 10 μm. **(B)** Schematic drawing of the TTN splicing reporter. **(C)** Ratio of Firefly luciferase to Renilla luciferase in HEK293 cells transfected with empty vector control, mouse RBM20, mouse RBM20-DupC, or both. **(D)** Ratio of Firefly luciferase to Renilla luciferase in HEK293 cells transfected with empty vector control, human RBM20, human RBM20-DupC, or both. ^****^ *p* value <0.0001. Significance was tested using a one-way ANOVA with Bonferroni post-hoc test.

### RBM20-DupC truncated protein does not have residual splicing activity nor does it act as a dominant negative

To check the splicing activity of RBM20-c.1222DupC, we used a previously reported splicing assay in HEK293 cells^22^. We overexpressed human and mouse WT RBM20 and RBM20-DupC in the presence of a TTN splicing reporter and measured the ratio of luciferase to renilla activity (Figure 3B). While WT RBM20 skipped the Fluc containing exon, RBM20-DupC was not sufficient to splice the reporter (Figure 3C-D). Additionally, upon transfecting the cells with increasing concentrations of RBM20-DupC, we observed no change in the splicing activity of the WT RBM20 (Figure 3C-D). This indicates that RBM20-DupC does not have a dominant negative effect on the splicing activity of the WT RBM20 protein.

### Healthy induced pluripotent stem cell derived cardiomyocytes engineered to carry RBM20-DupC display splicing abnormalities and mislocalization

To model the patient’s condition, we introduced the *RBM20* c.1222DupC mutation in a heterozygous manner into human induced pluripotent stem cells (iPSCs) from a healthy individual using prime editing (PE3). The presence of both the WT and DupC allele was confirmed in differentiated cardiomyocytes (Figure 4A-B). Immunostaining for α-actinin and troponin revealed a striated pattern in both WT and DupC+/– hiPSC-CM, consistent with proper sarcomeric organization and successful differentiation into cardiomyocytes (Figure 4C). Western blot analysis showed a ∼50% reduction in WT RBM20 protein in heterozygous cardiomyocytes compared with WT controls, while RBM20 mRNA levels remained unchanged (Figure 4D-E). This discrepancy suggests that the mutant transcript is not subjected to nonsense mediated decay (NMD), but instead that the c.1222DupC mutation may lead to reduced protein stability. We next assessed the splicing of known RBM20 targets. In DupC+/– hiPSC-CM, aberrant splicing was observed for titin (TTN) and RyR2 (Figure 4F). In addition, we measured expression of multiple cardiomyocyte markers, and found that the expression of GATA4 was similar between WT and DupC +/-hiPSC-CM, while Tnnt2 was slightly decreased (Figure 4G). Moreover, even though MYH7 is the dominant myosin protein in adult human cardiomyocytes, DupC+/– hiPSC-CM displayed a shift toward MYH6 expression (Figure 4H). Further transcriptome profiling by RNA sequencing revealed different clustering of both WT and DupC+/-reads and widespread gene expression changes in DupC +/-hiPSC-CM, with [1683] genes upregulated and [1,768] genes downregulated (|log2FC| > 0.5) (Figure 5A-B, Supplemental Table 3). KEGG pathway enrichment analysis showed that downregulated genes were enriched for categories related to hypertrophic cardiomyopathy, whereas upregulated genes were associated with extracellular matrix (ECM) receptor interactions and Wnt signaling (Figure 5C). Gene set enrichment analysis further revealed that downregulated transcripts in cellular component categories were enriched for genes involved in Z-disc organization, contractile fibers, and costameres, while upregulated transcripts included ECM- and collagen-related genes (Figure 5D, Supplemental Figure 2, and Supplementary Table 4). Terms related to Wnt signaling were also enriched in molecular function and biological process categories with decreased expression of genes involved in Lipid homeostasis (Supplemental Figure 2A-B). These results indicate that DupC +/-hiPSC-CM suggest a shift from a more contractile to a remodeling/fibrotic stage. We then used rMATS turbo, an established method for quantitative analysis of differential splicing events, to cluster splicing changes between WT and DupC +/-iPSC-CM, in five distinct categories: exon skipping (SE), intron retention (RI), mutually exclusive exons (MXE), and usage of alternative 3’ (A3SS) or alternative 5’ splice sites (A5SS). We used the reads on target and junction counts (JCEC) for each AS event. We observed changes for each of these splicing categories between the two cell populations, with the most extensive splicing changes affecting skipped exons (90 events in WT and 117 in DupC +/-) and mutually exclusive exons (10 events in WT and 21 in DupC +/-) (Figure 5E). ΔPSI (percent spliced-in) was calculated for the SE category to measure the exon inclusion differences between WT and DupC+/-(pvalue < 0.01 and ΔPSI > 0.1) (Supplemental Table 5). Among the top 50 differentially spliced skipped-exon events p<0.05, we detected multiple well-established RBM20 targets, including TTN, CAMK2D, and RyR2 (Figure 5F). Other splicing targets such as OBSCN, MTMR2 and CACNA1G also showed differential splicing (Supplementary Figure 2C). Overall, we demonstrate that the *RBM20* c.1222DupC mutation leads to missplicing of many known RBM20 targets.

**Figure 4.**
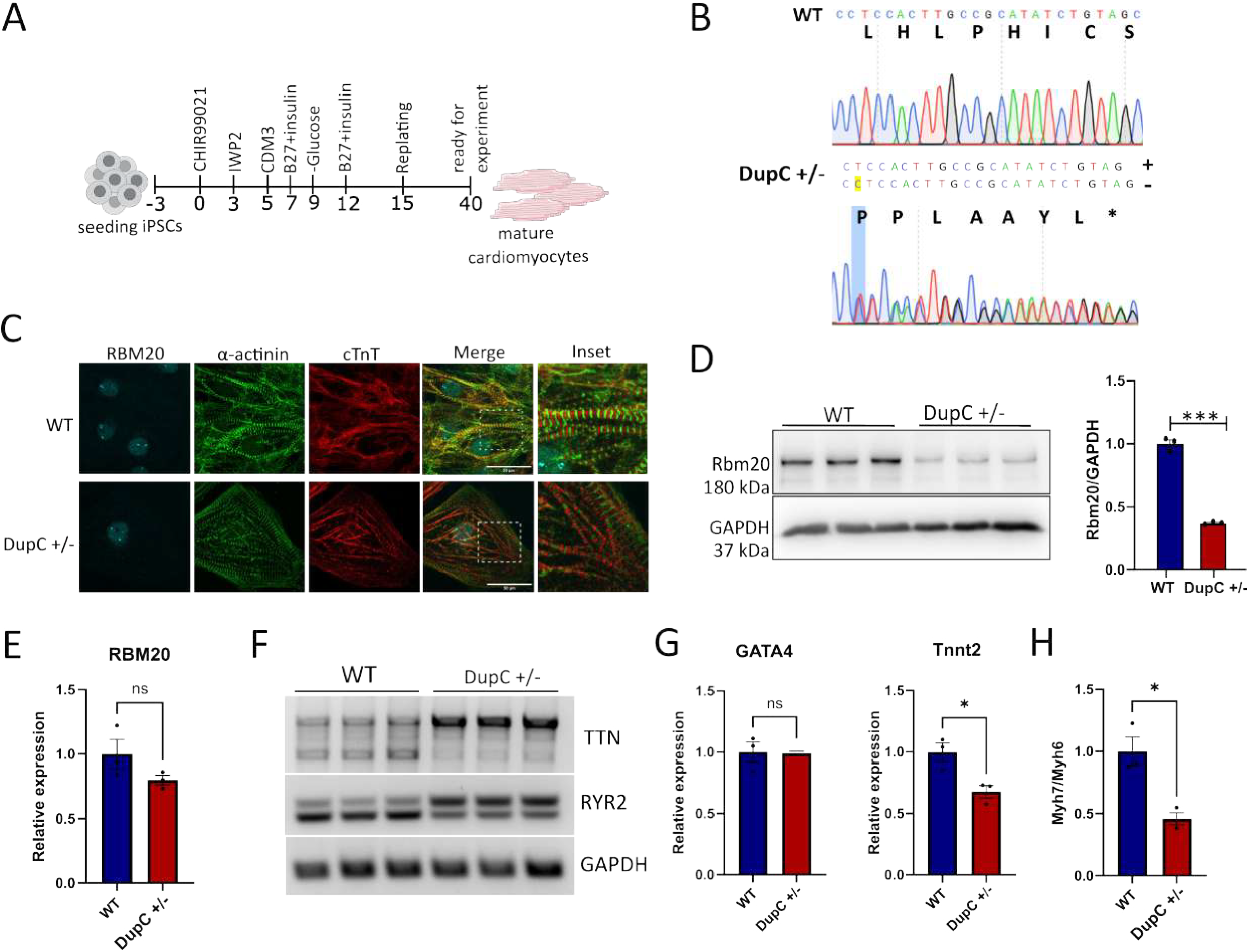
Generation of *RBM20* c.1222DupC hiPSC-CM. **(A)** Schematic representation of the workflow used to differentiate hiPSCs to cardiomyocytes. **(B)** Sanger sequencing of the RBM20-DupC (+/-) hiPSC-CM. In the DupC (+/-) panel, the top nucleotide sequence represents the WT sequence, and the bottom nucleotide sequence represents the mutated sequence with the duplicated C marked in yellow. Amino acid sequence is added underneath the nucleotide sequence. **(C)** Immunofluorescence of WT and Dupc (+/-) hiPSC-CM stained using anti-RBM20 (blue), anti-α-actinin (green), and cardiac Troponin T (red). Scale bar = 30 μm. **(D)** Immunoblotting of the cell lysate from WT and DupC (+/-) lines using anti-RBM20 antibody, the densitometric quantification presented as the bar graph **(E)** qPCR analysis of RBM20 in WT and DupC (+/-) iPSC-CM. **(F)** RT-PCR of RBM20 splice targets, Gapdh is used as control **(G)** qPCR analysis of Gata4 and cTnnt2 in WT and DupC (+/-) iPSC-CM. **(H)** Ratio of Myh7 to Myh6 in in WT and DupC (+/-) iPSC-CM. ^*^ *p* value < 0.05 ^**^ *p* value < 0.01, ^****^ *p* value <0.0001. Significance was tested using a two-tailed t-test.

**Figure 5.**
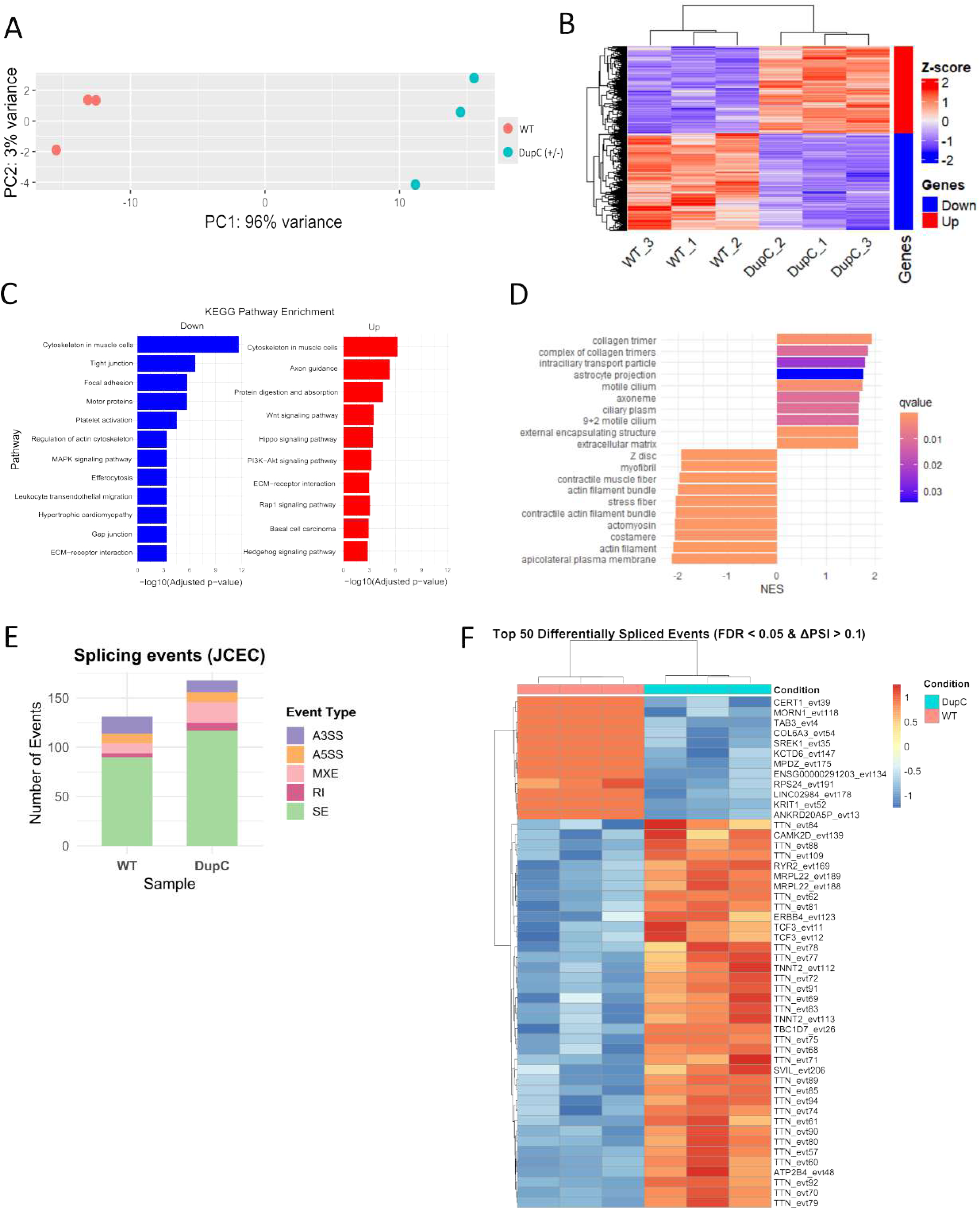
Transcriptomic analysis on *RBM20* c.1222DupC hiPSC-CM. **(A)** Principle Component analysis (PCA) plot of the reads from WT and DupC (+/-) iPSC-CM. **(B)** Heat map showing the differentially expressed genes (DEGs) in WT and DupC (+/-) iPSC-CM. **(C)** KEGG pathway enrichment analysis of the DEGs of DupC (+/-) iPSC-CM. **(D)** Gene set enrichment analysis (Cellular component) of the DEGs of DupC (+/-) iPSC-CM. **(E)** Total number of genome-wide differential splicing events (SE = exon skipping, RI = intron retention, MXE = mutually exclusive exons, A5SS = alternative 5’ starting site, A3SS = alternative 3’ starting site, events with an FDR < 0.01 and ΔPSI > 0.1 were included. **(F)** Heatmap of top 50 differential splicing events between WT and DupC (+/-) hiPSC-CM.

## DISCUSSION

Here, we report a novel heterozygous RBM20 mutation, specifically a duplication at nucleotide 1222 (c.1222DupC), in a patient with late-onset, mild dilated cardiomyopathy (DCM). This is distinct from most reported RBM20 mutations, which are predominantly missense single-nucleotide substitutions. Molecularly, overexpression of the mutant transcript produces a truncated protein that lacks splicing activity and does not exhibit a dominant negative effect on the WT RBM20 protein, suggesting that this mutation leads to haploinsufficiency. Most of the early reported mutations in RBM20 are located in the RS domain, and are associated with a more severe and early onset DCM phenotype. However, there are several reports now that show that mutations in other domains can also give rise to DCM. For example, patients with the V535I mutation in exon 6 within the RRM domain and the R716Q mutation located just outside the RS domain showed delayed development of DCM compared to the RS domain mutants^23^. Similarly, patients with the I536T mutation in the RRM domain encountered sudden cardiac death without any morphological changes and no known cardiac dysfunction^12^. This difference in disease severity between the RS-domain vs non-RS-domain mutations indicate that missplicing alone is not sufficient to fully explain the disease phenotype. Based on our overexpression and localization studies, we can conclude that *RBM20* c.1222DupC allele is capable of translation. However, we cannot confirm whether this truncated protein is stably expressed or detectable in patient tissue. The mutation introduces a premature termination codon, and it is possible that the majority of *RBM20* c.1222DupC transcripts undergo nonsense-mediated decay (NMD), thereby reducing RBM20 transcript and protein dosage *in vivo*. In contrast, our overexpression experiments did not reveal transcript reduction. However, these constructs contained only the coding sequence (CDS), and not the exon junctions, of RBM20 and would therefore bypass NMD. Importantly, in the *RBM20* c.1222DupC iPSC-CMs, we did not observe a decrease in the RBM20 mRNA level, which suggests that the mutant transcript is not subjected to NMD in a human cell model. Recently, Methawasin and colleagues showed that downregulating mutant RBM20 is beneficial in a mouse model with the RS-domain mutation RBM20-R639G^24^. However, our data shows that loss of RBM20 still leads to disease, which means that RBM20 downregulation as a therapeutic option should be very carefully evaluated. This is underscored by the fact that RBM20 KO models likewise have cardiac dysfunction^15,25^. RBM20 typically localizes to the nucleus, forming two characteristic puncta that correspond to the sites of TTN transcription which are crucial for RBM20 function, as the newly transcribed TTN pre-mRNA contains multiple RBM20 binding sites and acts as a scaffold for these RBM20 foci^26^. RBM20’s ability to localize to these foci is mediated by its RRM domain, and this spatial organization is thought to facilitate efficient RBM20-mediated splicing of its targets^26–28^. Furthermore, previous studies on RBM20 RS-domain mutants have demonstrated that the RS domain is essential for the proper nuclear localization of RBM20^24,7,8,30,31^. Variants in this domain inhibit the interaction on RBM20 with TNPO3, its nuclear transporter, and inefficient interaction between RBM20 and TNPO3, at least in part, responsible for this shuttling defect. Interestingly, when a nuclear localization signal is artificially introduced to an RS-domain variant, it restores normal splicing of RBM20 targets^7^. Given what is known about the function of these domains, it is surprising that the truncated RBM20-DupC protein is localized in both the nucleus and cytoplasm, even though it only contains the N-terminal leucine/proline-rich domain. Furthermore, a portion of the nuclear-localized DupC protein co-localizes with full-length RBM20 at the TTN transcription sites, albeit less efficiently than the WT protein. This raises important questions about the role of the N-terminal region of RBM20 in protein localization and function, as it appears to contain an uncharacterized mechanism for nuclear localization. In summary, we identify a novel truncating RBM20 mutation (c.1222DupC) that causes DCM through haploinsufficiency rather than through dominant-negative splicing defects. In addition, the unexpected nuclear localization of the truncated protein suggests the presence of an unrecognized function of the N-terminal leucine/proline-rich domain of RBM20. Together, these findings broaden the mechanistic spectrum of RBM20 cardiomyopathy and highlight dosage sensitivity as a critical disease determinant.

## Data Availability

All data produced in the present study are available upon reasonable request to the authors. RNA-sequencing data has been submitted to the GEO database and will be available upon publication of the manuscript.

## Acknowledgements

The authors gratefully acknowledge J. Fröhlich for technical help, and the data storage service SDS@hd supported by the Ministry of Science, Research and the Arts Baden-Württemberg (MWK) and the German Research Foundation (DFG) through grant INST 35/1503-1 FUGG.

## Dataset

The RNA-seq data generated in this study is submitted to the GEO database ……….

## Abbreviations

DCM: Dilated cardiomyopathy NMD Nonsense-mediated decay
NRCM: Neonatal rat cardiomyocytes
GSEA: Gene set enrichment analysis
iPSC-CM / hiPSC-CM: (Human) iPSC-derived cardiomyocyte
JCEC: Junction count Exon count
µm: micrometer

## List of Supplementary figures and tables

**Supplementary Figure 1.**
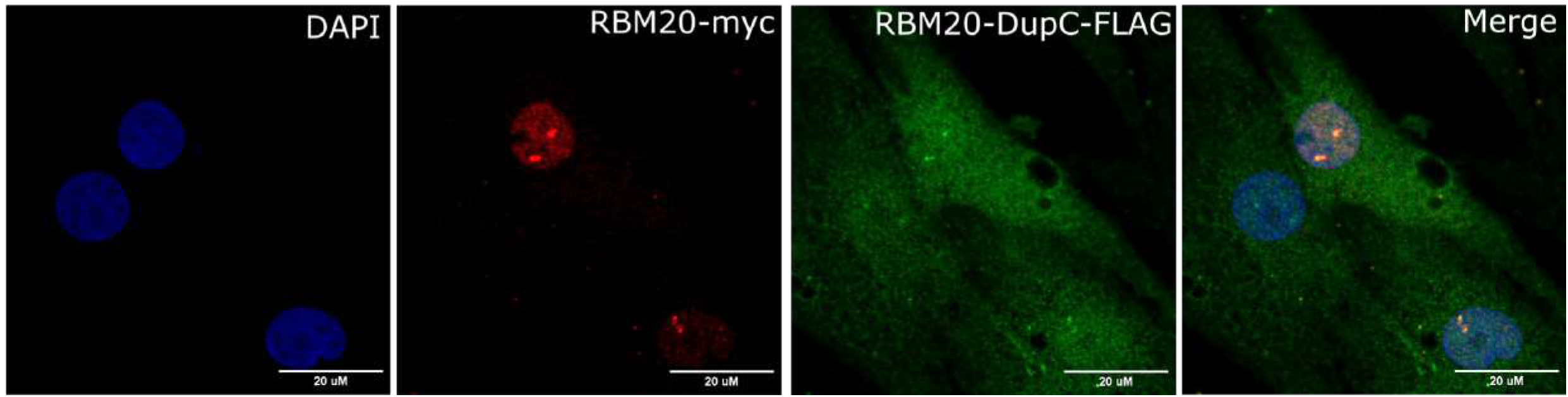
DupC partially colocalize with the full length RBM20. Immunofluorescence of NRCMs transfected with human myc-tagged RBM20 and FLAG-hRBM20-DupC. FLAG is stained in green, myc in red, and DAPI in blue. Scale bar is 20 μm.

**Supplementary Figure 2.**
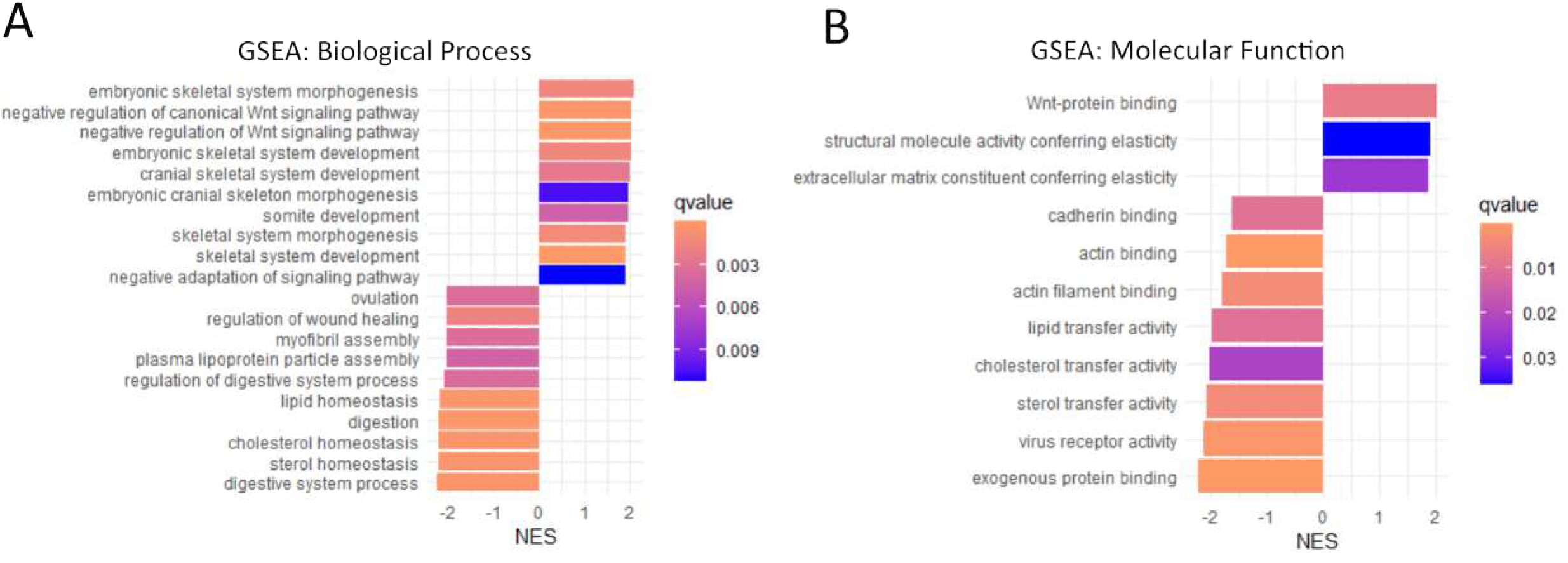
Transcriptomic analysis on *RBM20* c.1222DupC hiPSC-CM. **(A)** Gene set enrichment analysis (Biological process) of the DEGs of DupC (+/-) iPSC-CM. **(B)** Gene set enrichment analysis (Molecular Function) of the DEGs of DupC (+/-) iPSC-CM. **(C)** Heatmap of splicing events of known RBM20 targets between WT and DupC (+/-) hiPSC-CM.

**Supplemental Table 1** – List of oligos used for introducing c.1222DupC to iPSCs and oligos used for cloning and SDM of RBM20.

**Supplemental Table 2** - List of primers used for RT-PCR and qPCR.

**Supplemental Table 3** - List of differentially expressed genes in DupC (+/-) vs WT.

**Supplemental Table 4** - List of the enriched categories obtained after GSEA (Cellular component, Biological process, Molecular function).

**Supplemental Table 5** – List of alternative splicing events in DupC (+/-) vs WT.

